# *CYP2C19* Polymorphisms and Clinical Outcomes Following Percutaneous Coronary Intervention (PCI) in the Million Veterans Program

**DOI:** 10.1101/2023.10.25.23297578

**Authors:** Catherine Chanfreau-Coffinier, Kevin A. Friede, Mary E. Plomondon, Kyung Min Lee, Zhenyu Lu, Julie A. Lynch, Scott L. DuVall, Jason L. Vassy, Stephen W. Waldo, John H. Cleator, Thomas M. Maddox, Daniel J. Rader, Themistocles L. Assimes, Scott M. Damrauer, Philip S. Tsao, Kyong-Mi Chang, Deepak Voora, Jay Giri, Sony Tuteja, VA Million Veteran Program

## Abstract

**Background:** *CYP2C19* loss-of-function (LOF) alleles decrease the antiplatelet effect of clopidogrel following percutaneous coronary intervention (PCI) in patients presenting with acute coronary syndrome (ACS). The impact of genotype in stable ischemic heart disease (SIHD) is unclear.

**Objectives:** Determine the association of *CYP2C19* genotype with major adverse cardiac events (MACE) after PCI for ACS or SIHD.

**Methods:** Million Veterans Program (MVP) participants age <65 years with a PCI documented in the VA Clinical Assessment, Reporting and Tracking (CART) Program between 1/1/2009 to 9/30/2017, treated with clopidogrel were included. Time to MACE defined as the composite of all-cause death, stroke or myocardial infarction within 12 months following PCI.

**Results:** Among 4,461 Veterans (mean age 59.1 ± 5.1 years, 18% Black); 44% had ACS, 56% had SIHD and 29% carried a *CYP2C19* LOF allele. 301 patients (6.7%) experienced MACE while being treated with clopidogrel, 155 (7.9%) in the ACS group and 146 (5.9%) in the SIHD group. Overall, MACE was not significantly different between LOF carriers vs. noncarriers (adjusted hazard ratio [HR] 1.18, confidence interval [95%CI] 0.97-1.45, p=0.096). Among patients presenting with ACS, MACE risk in LOF carriers versus non-carriers was numerically higher (HR 1.30, 95%CI 0.98-1.73, p=0.067). There was no difference in MACE risk in patients with SIHD (HR 1.09, 95%CI 0.82-1.44; p=0.565).

**Conclusions:** *CYP2C19* LOF carriers presenting with ACS treated with clopidogrel following PCI experienced a numerically greater elevated risk of MACE events. *CYP2C19* LOF genotype is not associated with MACE among patients presenting with SIHD.

## Introduction

Combination therapy with a P2Y_12_ receptor inhibitor and aspirin is routinely prescribed to prevent major adverse cardiac events (MACE) in patients undergoing percutaneous coronary intervention (PCI).^1, 2^ Clopidogrel, a prodrug that is biotransformed into its active metabolite predominantly by the hepatic cytochrome P450 2C19 (CYP2C19) enzyme, is the most common antiplatelet drug prescribed in the setting of both acute coronary syndromes (ACS) and stable ischemic heart disease (SIHD).^3–5^ Approximately 30% of Whites and 60% of East Asians carry *CYP2C19* loss-of-function (LOF) alleles *(*2-*8)*, resulting in a decreased antiplatelet effect and an increased risk of MACE.^6–10^

Consensus guidelines published by the Clinical Pharmacogenetics Implementation Consortium (CPIC) recommend alternative antiplatelet therapy with prasugrel or ticagrelor in LOF carriers undergoing PCI as the effectiveness of these drugs is not impacted by the variants.^11^ These recommendations are not always followed due to the higher costs of the drugs and the increased risk of bleeding associated with the alternative agents.^4, 12–14^ However, the CPIC guidelines do not make a distinction between ACS and nonACS indications in their recommendations even though there are limited contemporary data evaluating the impact of *CYP2C19* variants in SIHD. Both observational and randomized controlled trial data show a strong association of *CYP2C19* LOF alleles and elevated MACE risk in patients treated with clopidogrel following PCI in patients with ACS.^9, 15–17^ This association has been conflicting or null in patients with SIHD.^18–20^

Additional data are needed in real-world cohorts of diverse patients that are not well represented in RCTs, including those with SIHD. The purpose of this study was to determine: 1) the association of *CYP2C19* LOF variants with MACE in participants receiving clopidogrel following PCI in the Veterans Health Administration (VA), 2) the association of the *CYP2C19* increased function variant with major bleeding events, and 3) whether these associations differed by indication for PCI (ACS vs. SIHD). The Million Veteran Program (MVP) is an ongoing, prospective genetic biorepository that recruits patients from 63 facilities within the VA, the largest integrated healthcare system in the US.^21^ The VA’s integration of longitudinal electronic health records (EHR) data with laboratory test results and pharmacy records makes the MVP an ideal platform for pharmacogenetic analyses.

## Methods

All relevant data are available in the main paper and supplemental data. Individual data cannot be shared publicly according to the Data Access Policy of the Million Veteran Program in the VA Office of R&D in Veterans Health Administration. The MVP received ethical and study protocol approval by the Veterans Affairs Central Institutional Review Board. Informed consent has been obtained from all participants. Each additional study was also approved by the local institutional review board. This project was also approved by the MVP Publication and Presentation Committee. For the full methods of this study, please refer to the Supplemental Methods.

## Results

### Patient characteristics

A total of 17,060 MVP participants underwent a PCI documented by the CART Program from 2009-2017. After applying the exclusion criteria, the final cohort was 4,461 (Supplemental figure 1). The cohort was 98% male, 76% white, with a mean age of 59.1 ±5.1 years (Table 1). Common comorbidities included hypertension (90%), dyslipidemias (86%), and diabetes (53%). The indication for PCI was ACS for 44% of the cohort and SIHD for 56%. The median follow-up time was 595 days (interquartile [IQR] range 584, 606) with no difference by PCI indication. The median (IQR) exposure to clopidogrel in the whole cohort was 383 days (301, 465) and was not different by PCI indication [ACS, 382 (305, 459) vs. SIHD, 383 (298, 468)].

**Table 1.**
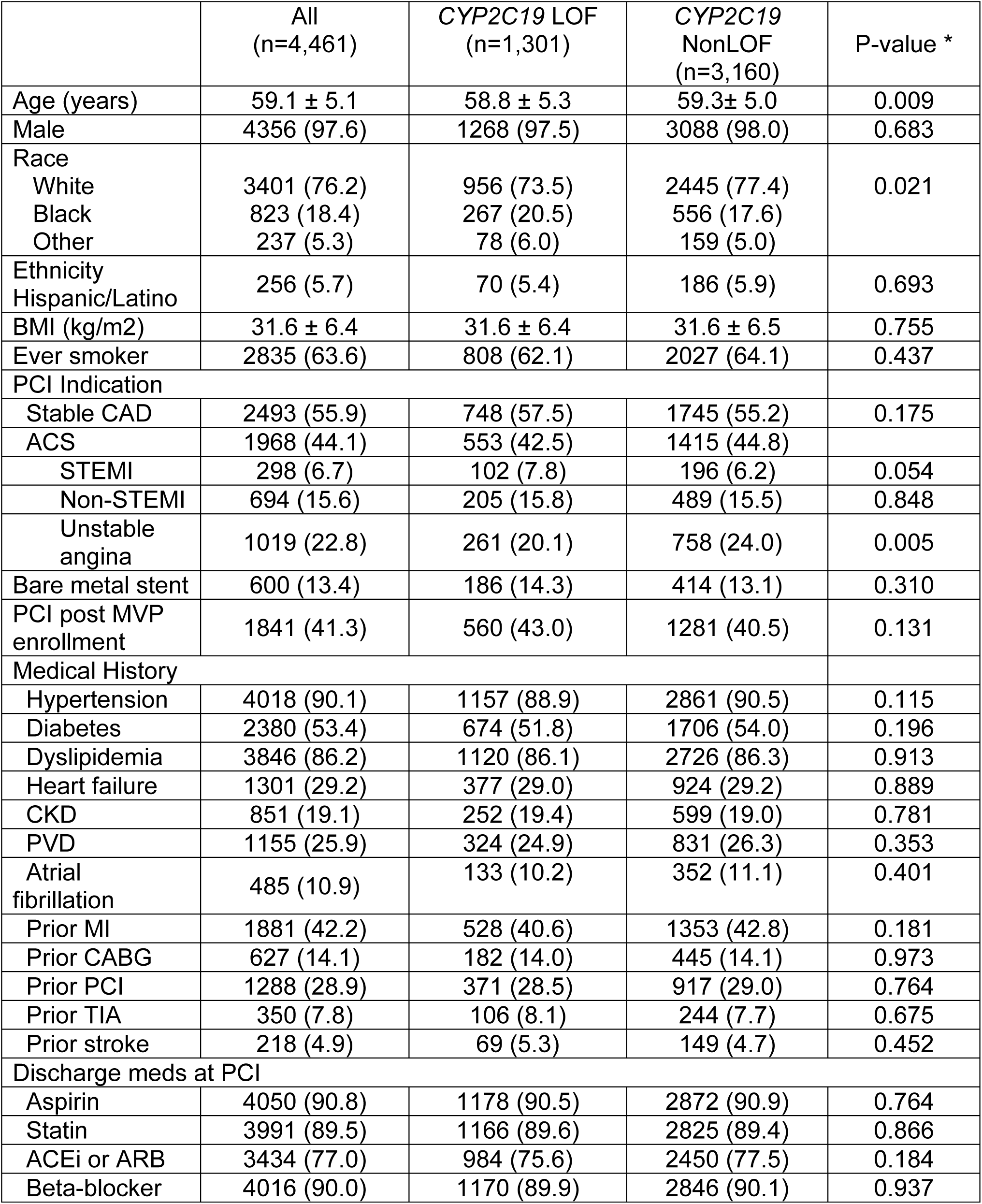
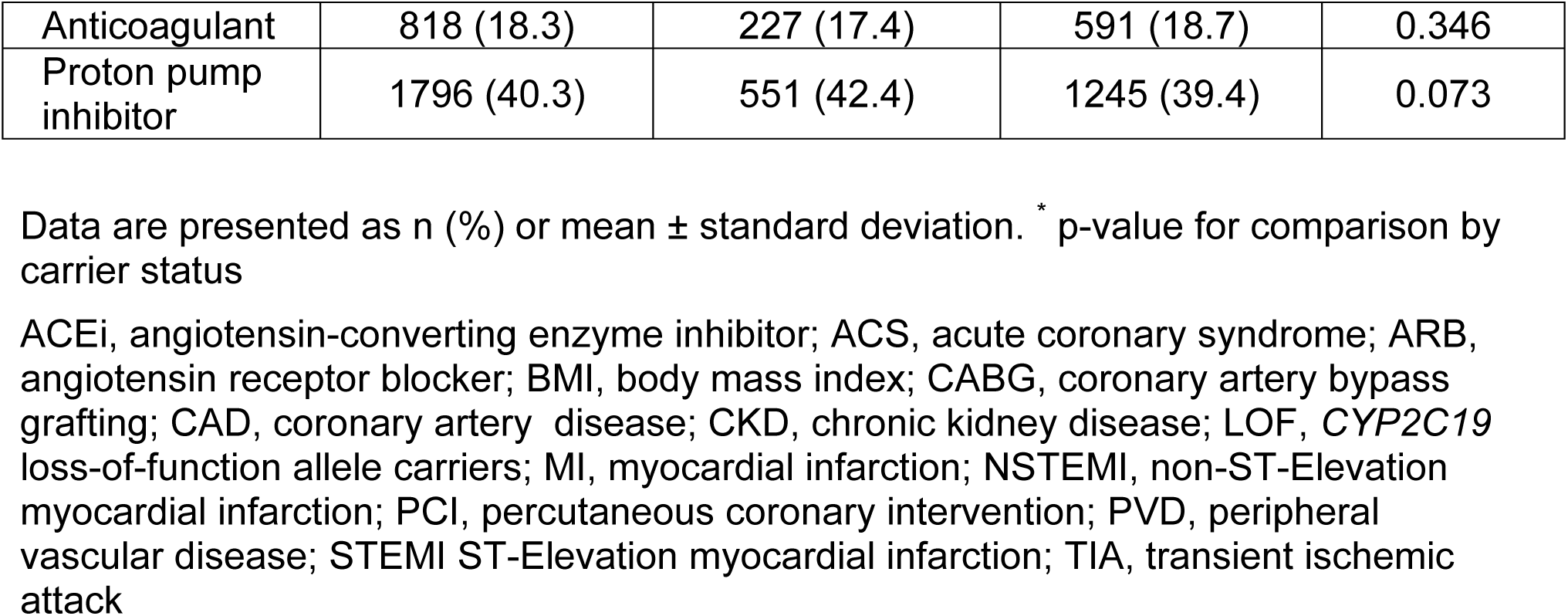
Baseline and procedural characteristics stratified by *CYP2C19* LOF carrier status.

|  | All<br>(n=4,461) | *CYP2C19* LOF<br>(n=1,301) | *CYP2C19*<br>NonLOF<br>(n=3,160) | P-value * |
| --- | --- | --- | --- | --- |
| Age (years) | 59.1 ± 5.1 | 58.8 ± 5.3 | 59.3 ± 5.0 | 0.009 |
| Male | 4356 (97.6) | 1268 (97.5) | 3088 (98.0) | 0.683 |
| Race |  |  |  |  |
| White | 3401 (76.2) | 956 (73.5) | 2445 (77.4) | 0.021 |
| Black | 823 (18.4) | 267 (20.5) | 556 (17.6) |  |
| Other | 237 (5.3) | 78 (6.0) | 159 (5.0) |  |
| Ethnicity |  |  |  |  |
| Hispanic/Latino | 256 (5.7) | 70 (5.4) | 186 (5.9) | 0.693 |
| BMI (kg/m<sup>2</sup>) | 31.6 ± 6.4 | 31.6 ± 6.4 | 31.6 ± 6.5 | 0.755 |
| Ever smoker | 2835 (63.6) | 808 (62.1) | 2027 (64.1) | 0.437 |
| PCI Indication |  |  |  |  |
| Stable CAD | 2493 (55.9) | 748 (57.5) | 1745 (55.2) | 0.175 |
| ACS | 1968 (44.1) | 553 (42.5) | 1415 (44.8) |  |
| STEMI | 298 (6.7) | 102 (7.8) | 196 (6.2) | 0.054 |
| Non-STEMI | 694 (15.6) | 205 (15.8) | 489 (15.5) | 0.848 |
| Unstable angina | 1019 (22.8) | 261 (20.1) | 758 (24.0) | 0.005 |
| Bare metal stent | 600 (13.4) | 186 (14.3) | 414 (13.1) | 0.310 |
| PCI post MVP enrollment | 1841 (41.3) | 560 (43.0) | 1281 (40.5) | 0.131 |
| Medical History |  |  |  |  |
| Hypertension | 4018 (90.1) | 1157 (88.9) | 2861 (90.5) | 0.115 |
| Diabetes | 2380 (53.4) | 674 (51.8) | 1706 (54.0) | 0.196 |
| Dyslipidemia | 3846 (86.2) | 1120 (86.1) | 2726 (86.3) | 0.913 |
| Heart failure | 1301 (29.2) | 377 (29.0) | 924 (29.2) | 0.889 |
| CKD | 851 (19.1) | 252 (19.4) | 599 (19.0) | 0.781 |
| PVD | 1155 (25.9) | 324 (24.9) | 831 (26.3) | 0.353 |
| Atrial fibrillation | 485 (10.9) | 133 (10.2) | 352 (11.1) | 0.401 |
| Prior MI | 1881 (42.2) | 528 (40.6) | 1353 (42.8) | 0.181 |
| Prior CABG | 627 (14.1) | 182 (14.0) | 445 (14.1) | 0.973 |
| Prior PCI | 1288 (28.9) | 371 (28.5) | 917 (29.0) | 0.764 |
| Prior TIA | 350 (7.8) | 106 (8.1) | 244 (7.7) | 0.675 |
| Prior stroke | 218 (4.9) | 69 (5.3) | 149 (4.7) | 0.452 |
| Discharge meds at PCI |  |  |  |  |
| Aspirin | 4050 (90.8) | 1178 (90.5) | 2872 (90.9) | 0.764 |
| Statin | 3991 (89.5) | 1166 (89.6) | 2825 (89.4) | 0.866 |
| ACEi or ARB | 3434 (77.0) | 984 (75.6) | 2450 (77.5) | 0.184 |
| Beta-blocker | 4016 (90.0) | 1170 (89.9) | 2846 (90.1) | 0.937 |
| Anticoagulant | 818 (18.3) | 227 (17.4) | 591 (18.7) | 0.346 |
| Proton pump inhibitor | 1796 (40.3) | 551 (42.4) | 1245 (39.4) | 0.073 |

Data are presented as n (%) or mean ± standard deviation. \* p-value for comparison by carrier status
ACEi, angiotensin-converting enzyme inhibitor; ACS, acute coronary syndrome; ARB, angiotensin receptor blocker; BMI, body mass index; CABG, coronary artery bypass grafting; CAD, coronary artery disease; CKD, chronic kidney disease; LOF, *CYP2C19* loss-of-function allele carriers; MI, myocardial infarction; NSTEMI, non-ST-Elevation myocardial infarction; PCI, percutaneous coronary intervention; PVD, peripheral vascular disease; STEMI ST-Elevation myocardial infarction; TIA, transient ischemic attack

Overall, 29% of the cohort (n=1,301) carried at least one loss-of-function (LOF) allele. Baseline comorbidities and discharge medications were well balanced between patients with and without LOF, although there were more Black participants among LOF carriers (Table 1). Baseline demographics by *CYP2C19* metabolizer groups, which included 1,743 NM (39%), 1,417 RM/UM (32%), 1,016 IM/PM (23%) and 285 IM/*17 (6%), are found in Supplemental table 5. Detailed breakdown of *CYP2C19* diplotypes by ancestry is shown in Supplemental table 6.

### Major Adverse Cardiovascular Events

In the overall cohort, 301 patients (6.7%) experienced MACE while being treated with clopidogrel, 155 (7.9%) in the ACS group and 146 (5.9%) in the SIHD group (Table 2, Supplemental table 7). The incidence of MACE in the overall cohort was not significantly different between LOF carriers and non-carriers (adjusted hazard ratio [aHR] 1.18, 95% confidence interval (CI) 0.97-1.45, p=0.096) (Table 2, Figure 1A, Supplemental figure 2) or by *CYP2C19* metabolizer group (Supplemental figure 3). However, ACS was strongly associated with MACE (Supplemental figure 2), and stratified analyses by PCI indication revealed a non-significantly higher risk of MACE in LOF carriers vs. non-carriers among patients with ACS (aHR 1.30, 95% CI 0.98-1.73; p=0.067, Table 2, Figure 1B, Supplemental figure 4). There was no impact of genotype in patients with SIHD (aHR 1.09, 95%CI 0.82-1.44, p=0.565) (Table 2, Figure 1C, Supplemental figure 5).

**Figure 1.**
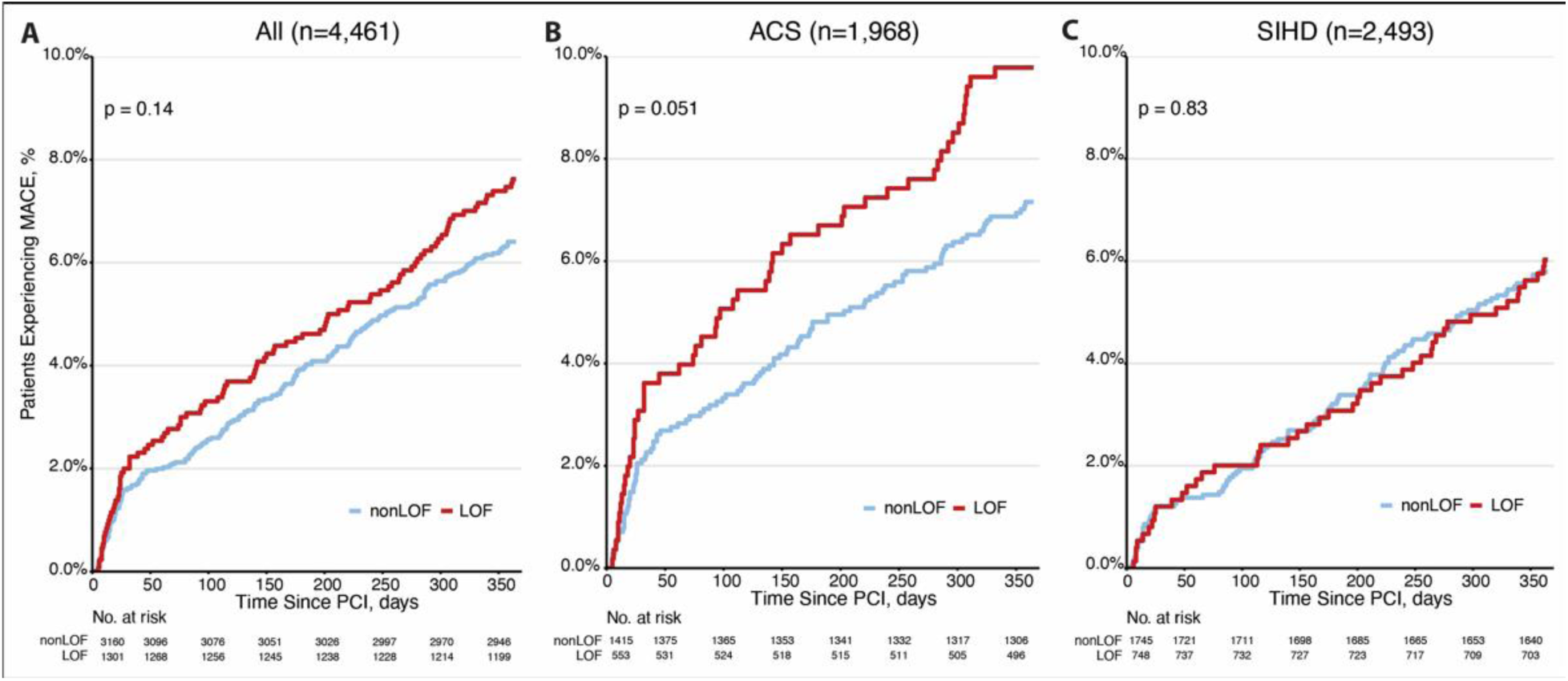
Time to major adverse cardiac event (MACE) in carriers of *CYP2C19* loss-of-function allele. A) in whole analytic sample (n=4,461), B) among patients presenting with acute coronary syndromes (ACS) (n=1,968), and C) among patients with stable ischemic heart disease (SIHD) (n=2,493). Cumulative incidence of MACE events using a Kaplan Meier model stratified by groups. MACE: major adverse cardiac event defined as myocardial infarction, stroke, and all-cause death; LOF: carrying at least 1 *CYP2C19* loss of function allele; nonLOF: without *CYP2C19* LOF alleles.

**Table 2.** Cox proportional hazard model for MACE in overall cohort and stratified by PCI indication.

| MACE outcome by *CYP2C19* LOF carrier status | Event rate | Unadjusted HR (95% confidence interval) | p-value | Adjusted HR (95% confidence interval) | p-value |
| --- | --- | --- | --- | --- | --- |
| Overall cohort (n=4,461)* |  |  |  |  |  |
| LOF (n=1,301) | 99 (7.6%) | 1.20 (0.94-1.52) | 0.141 | 1.18 (0.97-1.45) | 0.096 |
| nonLOF (n=3,160) | 202 (6.4%) | Reference |  | Reference |  |
| ACS (n=1,968)† |  |  |  |  |  |
| LOF (n=553) | 54 (9.8%) | 1.39 (0.998-1.93) | 0.052 | 1.30 (0.98-1.73) | 0.067 |
| nonLOF (n=1,415) | 101 (7.1%) | Reference |  | Reference |  |
| SIHD (n=2,493)† |  |  |  |  |  |
| LOF (n=748) | 45 (6.0%) | 1.04 (0.73-1.48) | 0.83 | 1.09 (0.82-1.44) | 0.565 |
| nonLOF (n=1,745) | 101 (5.8%) | Reference |  | Reference |  |

\*adjusted for indication for PCI (ACS vs. SIHD), age at PCI, race, use of bare metal stent, history of diabetes, chronic kidney disease, prior myocardial infarction, stroke, TIA, peripheral vascular disease, congestive heart failure, prior PCI, time of enrollment into MVP, with clopidogrel as a time varying exposure.
† adjusted for age at PCI, race, use of bare metal stent, history of diabetes, chronic kidney disease, prior myocardial infarction, stroke, TIA, peripheral vascular disease, congestive heart failure, prior PCI, time of enrollment into MVP, with clopidogrel as a time varying exposure.

### Bleeding events

At least one high risk factor for bleeding was found in 765 (17%) participants (Supplemental table 8), and 153 (3.4%) participants experienced a major bleeding event while on clopidogrel (Supplemental table 9). In the overall cohort we observed a non-significantly reduced rate of bleeding events among *CYP2C19* LOF carriers vs. non-carriers (aHR 0.73, 95% CI 0.50-1.4, p-value =0.096) (Table 3, Supplemental table 10. To investigate the contribution of *CYP2C19* increased function allele (*\*17*) towards bleeding events, we examined bleeding by *CYP2C19* metabolizer groups. The risk of bleeding was not increased among patients who carry at least one increased function copy [i.e., RM/UM and IM/*17 (Figure 2, Table 3)]. In contrast, the bleeding risk was significantly lower among IM/PM compared to NM (aHR 0.57, 95% CI 0.35-0.91, p-value =0.018, Supplemental figure 6).

**Figure 2.**
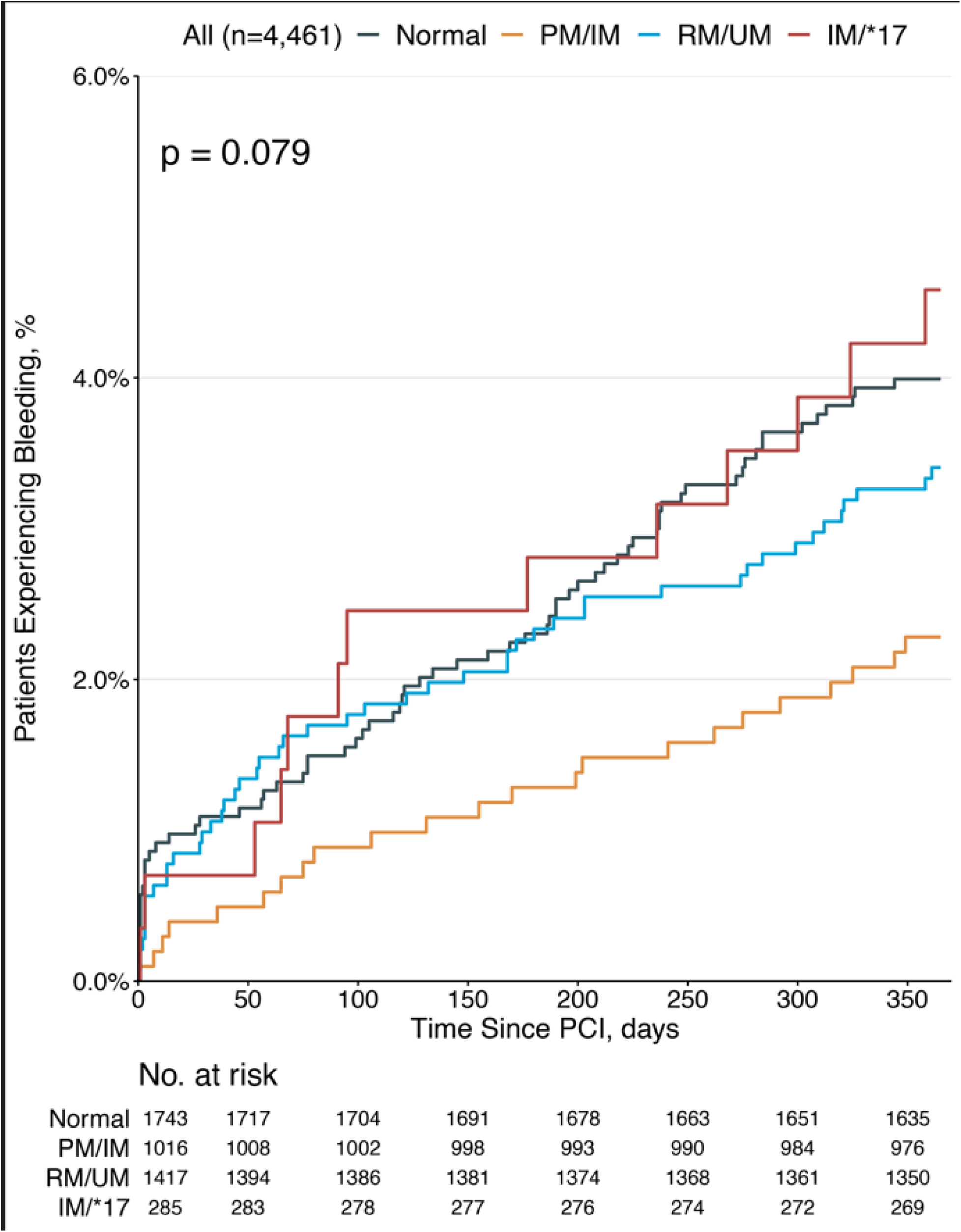
Time to bleeding event by *CYP2C19* metabolizer group in overall cohort (n=4,461). Cumulative incidence of bleeding events using a Kaplan Meier model stratified by groups. IM/PM: *CYP2C19* intermediate and poor metabolizers; RM/UM: *CYP2C19* rapid and ultra-rapid metabolizers; IM/*17: *CYP2C19* intermediate also carrying one or two copies of the *\*17* variant; NM: *CYP2C19* normal metabolizers.

**Table 3.**
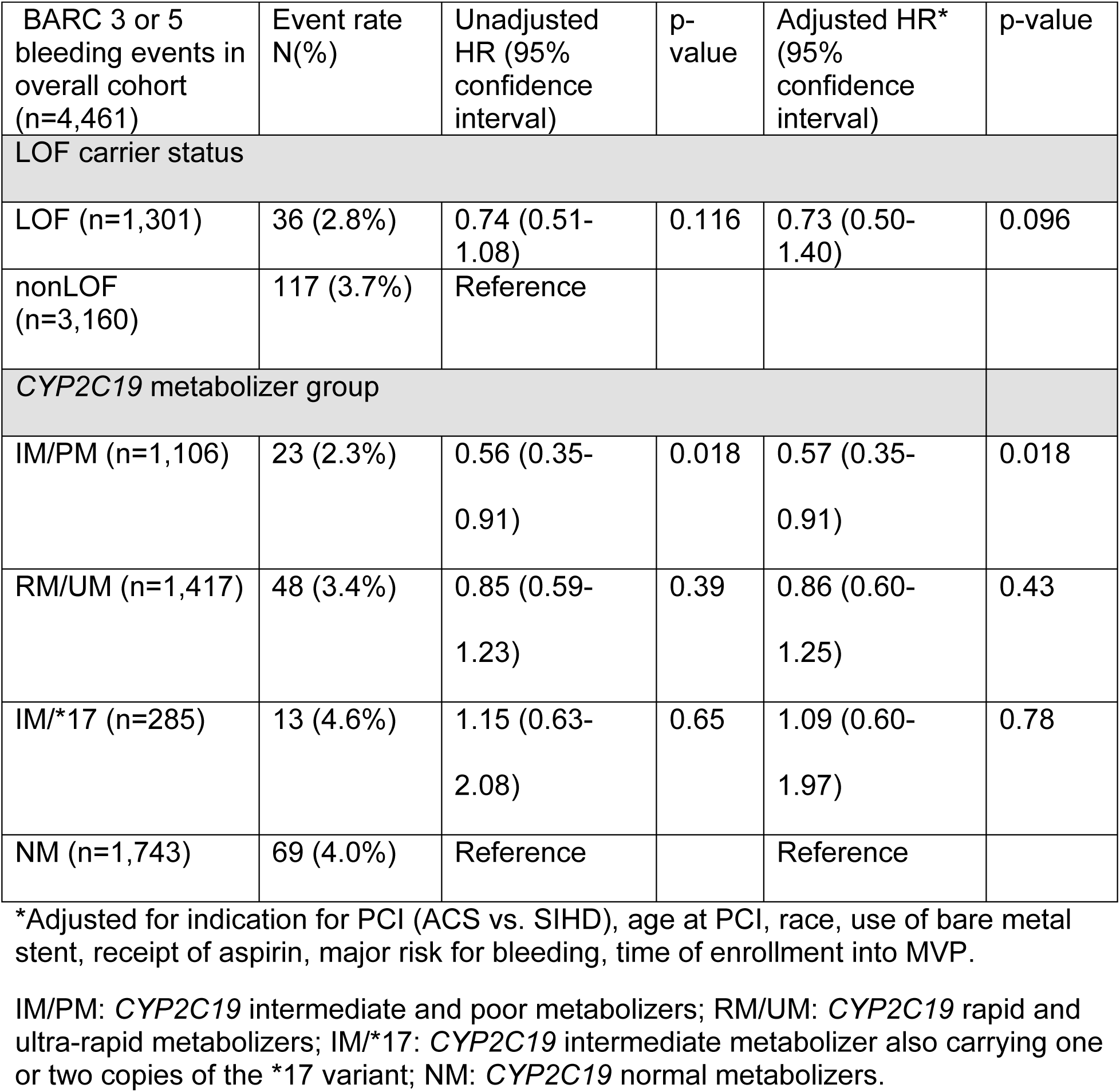
Cox proportional hazard model for bleeding events in overall cohort by *CYP2C19* loss of function carrier status and *CYP2C19* metabolizer group.

## Discussion

MVP participants age <65 years who were *CYP2C19* LOF carriers treated with clopidogrel following PCI did not experience a statistically increased risk of MACE. There was no impact of genotype in patients undergoing PCI for SIHD but among those with a PCI indication of ACS the risk of MACE demonstrated a numerically elevated, though marginally non-significant, 30% increased risk among LOF carriers versus noncarriers. The presence of the *CYP2C19* increased function *\*17* allele did not increase the risk of bleeding events, however LOF carrier status was associated with reduced risk of major bleeding events. These results have important implications for the role of genotyping to manage patients with stable coronary syndromes.

Our findings are largely consistent with previous observational studies, meta-analyses and recent prospective trials reporting an increased risk of MACE in *CYP2C19* LOF carriers treated with clopidogrel.^9, 17, 22–24^ The largest RCT, TAILOR-PCI, including 5,302 PCI patients, supported that prospective *CYP2C19* genotyping reduced the composite endpoint of CV death, MI, stroke, stent thrombosis and severe recurrent ischemia at 12 months in LOF carriers compared to LOF carriers in the control arm where all patients received clopidogrel (4.0% vs. 5.9%, respectively, [HR, 0.66; 95%CI, 0.43-1.02; p=0.06]). In a recent meta-analysis of 15,949 patients from 7 RCTs including TAILOR-PCI, predominantly focused on the ACS population (98%), the risk of ischemic events was significantly reduced in LOF carriers receiving ticagrelor or prasugrel (7.0%) versus clopidogrel (10.3%; HR, 0.70; 95%CI 0.59-0.83), while there was no such reduction in non-carriers (HR, 1.0; 95%CI 0.8-1.25). Most TAILOR-PCI participants presented with ACS with only 14% having SIHD and the authors did not examine the impact of genotyping in this subgroup.

A stratified analysis by PCI indication was conducted by Kim et al and found that patients with two *CYP2C19* LOF variants had a significantly higher risk of MACE in patients presenting with acute MI (hazard ratio, 2.88; 95% confidence interval, 1.27– 6.53; P=0.011).^18^ However, this finding was not seen in patients presenting with stable angina. A recent meta-analysis conducted to evaluate the association of *CYP2C19* LOF alleles and MACE risk in patients with SIHD and found an increased risk of MACE in LOF carriers.^19^ However, upon further inspection of the 21 studies they included in their analysis, eight studies also included patients with non-ST-elevation ACS (NSTE-ACS) and unstable angina (UA), and the indication for PCI could not be ascertained for three studies, limiting the overall conclusions of their analysis. More recently, an analysis by the IGNITE consortium in 3,342 patients, showed a decrease in MACE for LOF carriers presenting with ACS prescribed alternative therapy compared with LOF carriers prescribed clopidogrel (HR, 0.49; 95%CI 0.32-0.76, p=0.001) but no reduction in MACE in patients presenting with SIHD (HR, 1.02; 95%CI 0.44-2.36, p=0.96), indicating that prospective genotyping may be more important in patients undergoing PCI for ACS.^20^. In our analysis, the SIHD group included those without documentation of ACS (i.e., STEMI, NSTE-ACS, and UA). We found that the presence of *CYP2C19* LOF alleles in clopidogrel treated patients undergoing PCI for SIHD was not associated with an increased risk for MACE. These results support that a genotype-guided approach to antiplatelet prescribing may not be warranted in patients presenting with SIHD. In contrast to these previous studies, the definition of MACE in our study did not include repeat vascularization, an endpoint that is more prone to bias, and this conservative definition is therefore a strength of our study.

Our study included a more diverse real-world patient population with ∼20% African ancestry individuals and more patients with a higher rate of comorbid conditions. The clinical variables of age, BMI, CKD, diabetes along with *CYP2C19* genotype have been identified as independent predictors of high platelet reactivity on clopidogrel and adverse clinical outcomes.^25–27^ In comparison to TAILOR-PCI participants, those in MVP had a higher prevalence of these comorbidities including higher BMI (31.6 vs. 26.9 kg/m2), and more diabetes (53% vs. 28%) and CKD (19% vs. 12%). Future analyses in larger cohorts are warranted to evaluate the role of *CYP2C19* genotype in those with CKD or obesity/diabetes.

We did not find an association between the *CYP2C19 *17* allele and bleeding events. While several studies report an association of the *\*17* allele with decreased risk for MACE^28^ and/or increased risk of bleeding,^29^ this association is usually attenuated when accounting for the correlation due to linkage disequilibrium between the *\*2* and *\*17* alleles.^30^ While the presence of the *\*17* allele, by itself, is of limited utility in predicting MACE or bleeding events in patients receiving clopidogrel,^31, 32^ it may be useful as a component of a polygenic risk score to predict MACE.^33^ We also found a reduced rate of major bleeding among *CYP2C19* LOF carriers, which is likely related to diminished bioactivation of clopidogrel.

Several limitations to our analysis should be noted. Sample is limited to Veterans who received treatment within the VHA with a PCI documented in the CART data. We only included Veterans <65 years of age at the time of the procedure; this restriction aimed to address a possible lack in tracking clinical events in the VA electronic medical record for Veterans over age 65 who are eligible to receive care non-VA through Medicare. A gap in event documentation was supported by the observation of lower than expected frequency of MACE events among older patients (Supplemental Figure 7). As the risk of MACE increases with age^25–27^, a gap in capturing events in older patients would likely have abrogated the association of LOF alleles with the MACE outcome in the overall analyses. Future studies should aim to link Medicare and MVP data and document MACE occurring outside the VA among older patients. Despite the relatively large size of the sample, it is possible that the analyses lacked sufficient power enough to detect a significant association. We did not include a comparison group consisting of LOF carriers treated with prasugrel or ticagrelor, which would provide additional information about the potential reduction of MACE risk with these agents. At the time of cohort identification, there were few participants receiving these agents that also had genotype data available within MVP. For our bleeding analysis, we only counted severe bleeding events based on strict criteria including admission to the hospital or emergency department and objective measures of bleeding such as receipt of blood transfusion or defined changes in hemoglobin. However, patients frequently discontinue antiplatelet agents based on nuisance bleeding, which cannot be captured adequately using EHR data.

## Conclusions

MVP participants age <65 years who are *CYP2C19* LOF carriers treated with clopidogrel experienced a non-significantly higher risk of MACE when presenting with acute coronary syndromes. There was no impact of *CYP2C19* genotype in patients receiving PCI for stable ischemic heart disease. The presence of the *CYP2C19 *17* increased-function allele was not associated with an increased risk of bleeding events.

## Data Availability

All relevant data are available in the main paper and supplemental data. Individual data cannot be shared publicly according to the Data Access Policy of the Million Veteran Program in the VA Office of R&D in Veterans Health Administration.

## Disclaimer

This publication does not represent the views of the Department of Veterans Affairs or the United States Government.

## Abbreviation list

CART: Clinical Assessment, Reporting, and Tracking database
CYP2C19: Cytochrome P450 2C19
BARC: Bleeding Academic Research Consortium
LOF: Loss of function
MACE: Major adverse cardiac events
MVP: Million Veteran Program
PCI: Percutaneous coronary intervention
SIHD: Stable ischemic heart disease
STEMI: ST-segment elevation myocardial infarction

